# Who is more susceptible to Covid-19 infection and mortality in the States?

**DOI:** 10.1101/2020.05.01.20087403

**Authors:** Y Han, V.O.K. Li, J.C.K. Lam, P. Y Guo, R.Q Bai, W.T.W. Fok

**Affiliations:** Department of Electrical and Electronic Engineering, The University of Hong Kong, Pok Fu Lam, Hong Kong; CEEPR, MIT Energy Initiative, MIT, Cambridge, Massachusetts, USA; Department of Computer Science and Technology, The University of Cambridge, Cambridge, UK; Clare Hall, The University of Cambridge, UK

**Keywords:** Covid-19, infection, mortality, socio-economic status (SES), social distancing, lockdown, county/state-level, US

## Abstract

**Background:** A novel coronavirus was detected in Wuhan, China and reported to WHO on 31 December 2019. WHO declared a global pandemic on 11 March 2020. The first case in the US was reported in January 2020. Since mid-March 2020, the number of confirmed cases has increased exponentially in the States, with 1.1 million confirmed cases, and 57.4 thousand deaths as of 30 April 2020. Even though some believe that this new lethal coronavirus does not show any partiality to the rich, previous epidemiological studies find that the poor in the US are more susceptible to the epidemics due to their limited access to preventive measures and crowded living conditions. In this study, we postulate that the rich is more susceptible to Covid-19 infection during the early stage before social distancing measures have been introduced. This may be attributed to the higher mobility (both inter- and intra-city), given their higher tendency to travel for business/education, and to more social interactions. However, we postulate after the lockdown/social distancing has been imposed, the infection among the rich may be reduced due to better living conditions. Further, the rich may be able to afford better medical treatment once infected, hence a relatively lower mortality. In contrast, without proper medical insurance coverage, the poor may be prevented from receiving timely and proper medical treatment, hence a higher mortality.

**Method:** We will collect the number of confirmed Covid-19 cases in the US during the period of Jan 2020 to Apr 2020 from Johns Hopkins University, also the number of Covid-19 tests in the US from the health departments across the States. County-level socio-economic status (SES) including age, sex, race/ethnicity, income, education, occupation, employment status, immigration status, and housing price, will be collected from the US Census Bureau. State/county-level health conditions including the prevalence of chronic diseases will be collected from the US CDC. State/county-level movement data including international and domestic flights will be collected from the US Bureau of Transportation Statistics. We will also collect the periods of lockdown/social distancing. Regression models are constructed to examine the relationship between SES, and Covid-19 infection and mortality at the state/county-level before and after lockdown/social distancing, while accounting for Covid-19 testing capacities and co-morbidities.

**Expected Findings:** We expect that there is a positive correlation between Covid-19 infection and SES at the state/county-level in the US before social distancing. In addition, we expect a negative correlation between Covid-19 mortality and SES.

## Significance Statement

To the best of our knowledge, this is the first study that examines the relationship between SES and Covid-19 infection and mortality at fine-grained spatial resolution (state/county-level) across the US, while accounting for the effects of lockdown/social distancing, comorbidities and the limitations of Covid-19 testing capacities.

## Introduction

Covid-19 is a highly contagious disease. A report published by the CDC in the US suggested that the median R0 value was 5.7 during the early outbreak in Wuhan, China (1). The first Covid-19 infection case was reported in the US in Jan 2020. Since late March, the total number of confirmed infections and mortalities in US has superseded other countries in the world.

The 1918 Spanish flu pandemic suggests that socio-economic factors can affect the mortality rate, though their effects on the infection rate is minimal. Although it is generally believed that the chance of getting an infectious disease and dying from it is equal across different socio-economic status (SES) groups (2), epidemiological studies have suggested that the risks of mortality may be unevenly distributed across these groups (3, 4). People having a lower SES in the States are more susceptible to Covid-19 infection due to much more limited access to preventive care and poorer living conditions. The pandemic is a new and highly contagious respiratory disease; no effective medical prevention/treatment is currently available to contain the problem. Despite how Covid-19 infection and mortality vary across different SES groups in the US remains largely unknown, such variations will have significant implications on public health.

We hypothesize that at the beginning, a higher SES will suffer a higher infection rate due to the following reasons:

1. People of a higher SES travel more: Surveys have unequivocally shown that the frequency of travel is correlated with income. A survey of 6 million people in 25 cities conducted by VISA group has confirmed that the fraction of people who undertake international flights increases with the increase in annual incomes, irrespective of age (5). Another survey by Ipsos has also found that the number of airline trips increases with an increase in annual household incomes in general (6).
2. People of a higher SES would engage in more social contacts: Literature reveals that people of a higher SES generally are more inclined to engage in group activities (organized by churches or NGOs) and socialize with others. Saffer (2005) also finds that an increase in incomes generally increases the number of memberships of and visits to social organizations (7). Brehm and Rahn (1997) notes that the real family income is an important determinant of group activities (8). Alesina and La Ferrara (2002) claims that lower incomes reduce social trusts (9). Leung et al. (2017) reports that both the average number of contacts and contact hours per day increase as the income level increases (10). In addition, surveys based on large, nationally representative samples of Americans (N=118,026) show that people earning higher incomes spend less time with their families and neighbours and more time with their friends (11).
3. People of a higher SES tend to send their kids to travel/study in other states or countries; Evidence shows that students from a higher income family are more likely to study abroad. Open door survey finds that the majority (70%) of the US students studying abroad are white(12). Dessoff (2006) notes that for the working-class, sending their children to study abroad may seem an unreachable task (13). Salisbury et al. (2008) finds that household income is an important factor determining whether students would choose to study abroad (14). A higher income also leads to a higher chance of enrolling in out-of-state colleges. Curs and Singell (2002) finds that students from a well-off family tend to attend out-of-state colleges than those from a lower income family (15). Perna and Titus (2004) finds that fewer low-SES high school students (5%) have enrolled in out-of-state institutions as compared to high-SES counterparts (31%) (16).

We hypothesize, once infected, people with a higher SES will have a lower mortality with better healthcare.

### Factors affecting Covid-19 infection and mortality

A systematic review is conducted based on the Covid-19 Open Research Dataset (CORD-19), which covers more than 38,000 academic articles on Covid-19 and other zoonotic coronaviruses such as SARS and MERS. The related factors of Covid-19 infection and mortality are identified as follows:

- Demographic: age, sex (17)
- Medical condition: co-morbidity (18)
- Lifestyle: smoking (19)
- Environment: weather (20), air pollution (21)
- Public health intervention: lockdown/social distancing (22)

To date, in the US, the role of socio-economic status (SES) in Covid-19 infection and mortality is yet to be fully investigated. Previous studies have shown that SES could be linked to influenza infection and mortality in the US (3, 4). Recent evidence in New York City suggests that people with a lower SES is more likely to get infected, though the correlation is relatively weak (23). Some recent US news articles hypothesize that SES could affect Covid-19 infection (24) and/or mortality in the country (24, 25). (24) suggests that the poor is more likely to get infected and die from Covid-19, based on previous epidemic experience. However, such arguments are yet to be substantiated by the existing Covid-19 evidence. (25) reveals the relationship between race and Covid-19 infection/mortality. Based on the US Covid-19 Map and Census data, the Washington Post shows how infection and mortality cases are unequally distributed among different racial groups, with more blacks and Hispanics suffering from Covid-19, in some parts of the country.

Up till now, no rigorous scientific study has been done to investigate the role of SES in Covid-19 infection and mortality across the US, at the fine-grained spatial resolution. To fill this research gap, we propose to examine the relationship between SES and Covid-19 infection/morality at the state/county-level in the US, while accounting for other important factors such as lockdown/social distancing, co-morbidities, and COVID-19 infection testing capacities.

## Result

We expect there being a positive correlation between Covid-19 infection and SES at the state/county-level in the US before social distancing. In addition, we expect a negative correlation between the Covid-19 mortality and the SES.

## Discussion

### Research significance

To the best of our knowledge, this is the first study that investigates the relationship between SES and Covid-19 infection/morality at the state/county-level across the US.

In our regression analysis, we have accounted for the effects of social distancing/lockdown, co-morbidities, and the limitations of COVID-19 testing capacities.

### Implication

We expect that in the States, the rich are at a higher risk of Covid-19 infection before social distancing/lockdown, but a lower risk of mortality, after controlling other confounding factors, such as social distancing/lockdown, co-morbidities, and the limitations of COVID-19 testing capacities.

We expect that in the US, while the rich may have a higher risk of Covid-19 infection before social distancing/lockdown, the poorer may have higher mortality after being infected. This can help guide future public health policy-making, as identifying which particular income groups are most/least susceptible to Covid-19 infection/mortality, should provide insights into how future medical resource should be allocated or redirected, in order to help the most needy in the society.

We expect that in the future, studies on Covid-19 infection/mortality modelling should incorporate SES factors which should have a decisive impact on the accuracy of such modeling.

## Material and Method

### Unit of Analysis and Data Collection

Our study covers the period from 20 January 2020 (when the first case was reported in the US) to 30 April 2020 (26). States having the highest number of confirmed cases per million population are selected, including New York, New Jersey, Louisiana, Massachusetts, and Connecticut etc.

Data collected on a daily basis at the state/county-level. The number of confirmed cases/deaths are aggregated as the cumulative number of confirmed cases/deaths before and after lockdowns at the state/county-level.

### Data Source

Covid-19 infection and mortality can be represented by the daily number of confirmed cases and deaths. They are collected from Johns Hopkins University’s Covid-19 Map which obtains its data from the US CDC. Covid-19 testing capacity is the daily number of people who get tested. The data can be obtained from the COVID Tracking Project which collects data from the health departments of different states (27). SES can be represented by population size/density, age, sex, race/ethnicity, income, education, occupation, employment status, immigration status, and housing price. The data are collected from the US Census Bureau. Co-morbidities to Covid-19 are represented by the incidence rates of chronic diseases, including diabetes, COPD, asthma, cancer, and hypertension. The data are collected from the US CDC.

Lockdown/social distancing refers to the state/county-level policies (in particular, stay-at-home orders) imposed by the state/county-level governments. The data are obtained from the state/county government websites (28). Mobility/movement is indicated by the inter-country movement, represented by the number of international flights between the subject states/counties and the countries of high COVID-risks (European countries and China), as well as the intra-country movement, represented by the number of domestic flights between the subject states/counties and other subject states/counties.

### Statistical Analysis

Two regression models are used to examine the relationship between the SES and the Covid-19 infection, and between SES and Covid-19 mortality, before and after lockdown. The following SES indicators are adopted:

- Household income (4)
- Other SES variables will be included whenever relevant, such as education, immigration status, housing price, and employment status (29)
- Need to account for the co-linearity among the SES variables (30)

Dependent variables include cumulative number of confirmed cases/deaths before and after the lockdown policy has been introduced. Independent variables include social distancing/lockdown, COVID-19 testing capacity, SES, co-morbidity, mobility.

## Data Availability

The data is available from the corresponding authors upon reasonable request.

